# BPF-GILD study: an observational cohort study of UK pigeon fanciers

**DOI:** 10.1101/2024.07.27.24311102

**Authors:** Richard J Allen, Jude Wellens-Mensah, Olivia C Leavy, Stephen Bourke, Will Henderson, Howard Smith, Emma Johnson, Daniella Marks, Ye Myat, Cecillia Rafique, Gina Parcesepe, Tamara Hernandez-Beeftink, Beatriz Guillen-Guio, Eric Cheng, Carmen Chan, Gabrielle Clark, Stuart Dickson, Andrew Pearson, Mathew Embley, Phillip Lynch, Gavin Boyd, Bibek Gooptu, Yuan Ji, Charles McSharry, Louise V Wain, Mark Spears

## Abstract

**Introduction:** Hypersensitivity pneumonitis (HP), a common interstitial lung disease (ILD), comprises two distinct but related forms, acute and fibrotic (fHP). HP is classically described as a disease triggered by antigen exposure. However, a wide range of triggers are described and in ∼50% of cases no cause is identified, rendering observational studies challenging. The British Pigeon Fanciers Genetics of ILD (BPF-GILD) study aims to address this by studying a population with a clear history of exposure to a common trigger.

**Methods:** Participants were recruited from 2019 to 2023 at large UK Pigeon Fancier meetings. Each participant performed spirometry, completed a standardised questionnaire with a doctor, and provided blood samples. We present our baseline data in this manuscript.

**Results:** 417 subjects were recruited from four shows. The median age of the cohort was 63 years, 95% were male and 94% self-reported white ethnicity. The median number of pigeons kept was 80 [range 4-800], with fanciers spending 14 hours per week [1-100] in their lofts. 52% of participants had occupational dust exposures.

49% of the cohort reported at least one respiratory symptom related to loft exposure. 14% had a history of ILD and these individuals had more loft-related respiratory symptoms, poorer lung function, and appeared more likely to wear a mask with their pigeons than those without (74% vs 57%). 41% of participants had positive responses to questions employed to detect occult connective tissue disease in ILD clinics.

**Discussion:** Our well characterised cohort of pigeon fanciers commonly experience acute HP symptoms and are likely to be at increased risk of fHP. Subsequent work using stored samples will enable us to determine genetic risk factors and pathways relevant to the development of fHP.

## Introduction

Hypersensitivity pneumonitis (HP), a common form of interstitial lung disease, is classified into acute and fibrotic forms (1). HP is classically described as an aberrant response to environmental exposures. However, only a small proportion of subjects develop HP after exposure to relevant precipitants, likely reflecting variation in immune responses and underlying genetic risk (1–4).

Acute HP, which classically presents as a flu-like illness several hours after exposure, is largely spontaneously reversible, with risk of recurrence managed by reducing exposure to the trigger (1,4,5). However, whilst acute HP is relatively well studied (1), the factors driving the development of fibrotic HP (fHP) are unclear despite the recognised link between the two forms of the condition (1,5). Progress in the study of fHP has arguably been impaired by challenges in diagnosis, mainly related to distinction from other progressive fibrotic interstitial lung diseases (ILDs), particularly idiopathic pulmonary fibrosis (IPF) which shares radiologic similarities (6). Therefore improvements in our understanding of the key steps for the development of fHP are urgently required, as this would allow the development of specific non-invasive diagnostic tests and assessment of treatments specific to patients with HP.

Whilst multiple different environmental exposures have been linked to the development of HP (1), exposure to pigeons is amongst the most common and well-documented. Pigeon fancying is a well-established sport within the UK, with the Royal Pigeon Racing Association (RPRA) having 22,500 members in 2016. Active fanciers are known to be at increased risk of acute and fHP, and other ILDs, compared to matched subjects (7). UK pigeon fanciers therefore provide a population of individuals with known and consistent exposure to a well described cause for HP.

The British Pigeon Fanciers Genetics of ILD (BPF-GILD) study, which commenced in January 2019, aims to detect genetic and immunological factors of relevance to the development, diagnosis and management of HP. BPF-GILD builds on a long history of ‘pigeon lung’ research supported by the RPRA, and led by the pigeon fancying community, in collaboration with clinicians and scientists since 1967 (ref: https://www.rpra.org/wp-content/uploads/2012/03/pigeon-fanciers-lung-medical-team-report-2015.pdf).

Herein we report baseline characteristics of the individuals recruited to the BPF-GILD study up to and including the January 2023 RPRA meeting.

## Methods

Participants were recruited during their attendance at large national and regional meetings of the RPRA and Scottish Homing Union (SHU). The annual RPRA 2-day ‘Show of the Year’ in Blackpool is attended by up to 20,000 pigeon fanciers, whilst the annual SHU in Lanark attracts around 400 attendees.

Ethical approval was provided by North West - Greater Manchester West Research Ethics Committee (18/NW/0843) in December 2018.

To be eligible, participants had to be active in the sport, with ongoing regular exposure to pigeons at the time of recruitment. Once consented, each participant provided information to allow completion of a questionnaire by an experienced clinician (**Supplement**). This collected information about current symptoms, to allow allocation of a modified MRC dyspnoea (mMRC) score, current diagnosis of interstitial lung disease or connective tissue disease, demographic characteristics (including weight and height), smoking history (including pack-years smoked and smoking cessation duration where applicable) and occupational dust exposures. In addition, a series of questions utilised in ILD clinics to identify symptoms suggestive of an occult connective tissue disease (subsequently described as ‘CTD symptoms’, see **Supplementary Methods**) were employed. Participants were also asked if they had experienced any eye, upper airway, respiratory or systemic symptoms related to their exposure to pigeons (see **Supplementary Methods**). Finally, information was sought on length of time in the sport, number of pigeons kept, and other pigeon keeping habits. Individuals were defined as having a “suspected ILD” if they answered yes to having pulmonary fibrosis or been diagnosed by a doctor with ‘pigeon lung’ (where this was associated with their having attended a hospital clinic and a subsequent chest CT scan).

Subjects performed lung function testing, under the guidance and supervision of pulmonary physiologists, or trained doctors. The majority performed spirometry alone (obtained using EasyOn PC Spirometer, NND Technologies), with subsets also completing transfer factor (EasyOne Pro, NND Technologies) and/or forced oscillometry (Resmon Pro device, MGC Diagnostics) measurements. Spirometry results obtained from tests of quality grade D or above (8) were included in the analysis. Blood samples were collected for DNA, RNA, serum proteomics, specific IgG to pigeon antigens, and other immunological tests (**Supplementary Methods**). Pigeon specific IgG results were returned to participants by post within 3 months of their being obtained.

Data generated was processed for presentation using R v4.0.1. No significance tests have been performed.

## Results

Data from recruitment at four events (RPRA ‘Show of the Year’ in Blackpool January 2019, 2020 and 2023, and SHU annual meeting in Lanark in December 2019) are presented (**Supplementary Figure 1**). No recruitment took place in 2021 or 2022 due to restrictions related to the COVID-19 pandemic.

Four hundred and seventeen individuals consented to participate by the end of the recruitment event in January 2023, of which four hundred and thirteen had data available from either a questionnaire or spirometry. Thirty participants had requested withdrawal from further contact by the study at the end of January 2023. Of these two had died during follow up, and one withdrew consent for ongoing follow up due to his wife developing HP resulting in his ceasing to keep pigeons. The remainder did not volunteer a reason for withdrawal. Sixty seven participants attended for repeat visits, with twenty six attending on three occasions.

### Baseline participant characteristics

Basic demographics and pigeon keeping activity are reported in Table 1. The majority of study participants were male (95%) and of white ethnicity (94%). The median age of participants was 63 years (Supplementary Figure 2). Of the three hundred and eighty individuals with BMI recorded, three hundred and thirty-two had a BMI≥25 (classified as overweight) and one hundred and eighty had a BMI ≥30 (classified as obese).

**Table 1:**
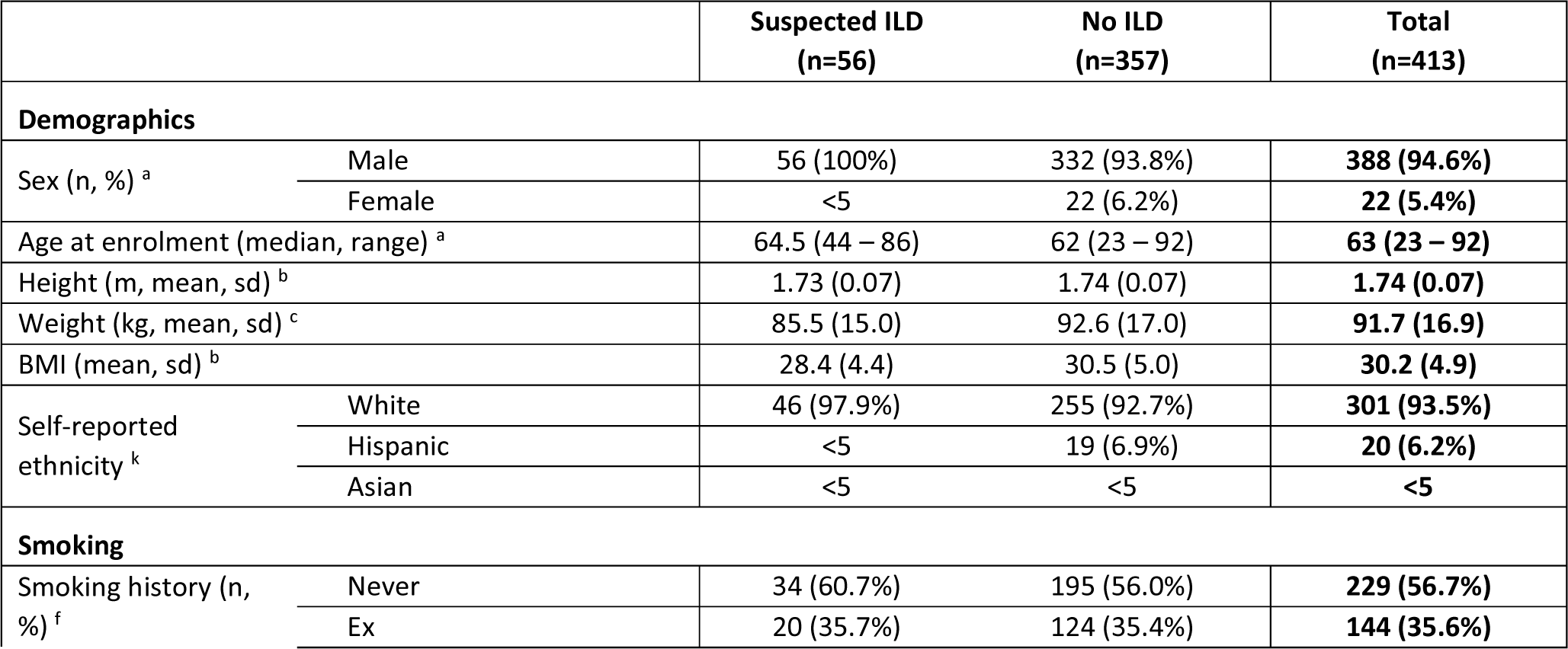
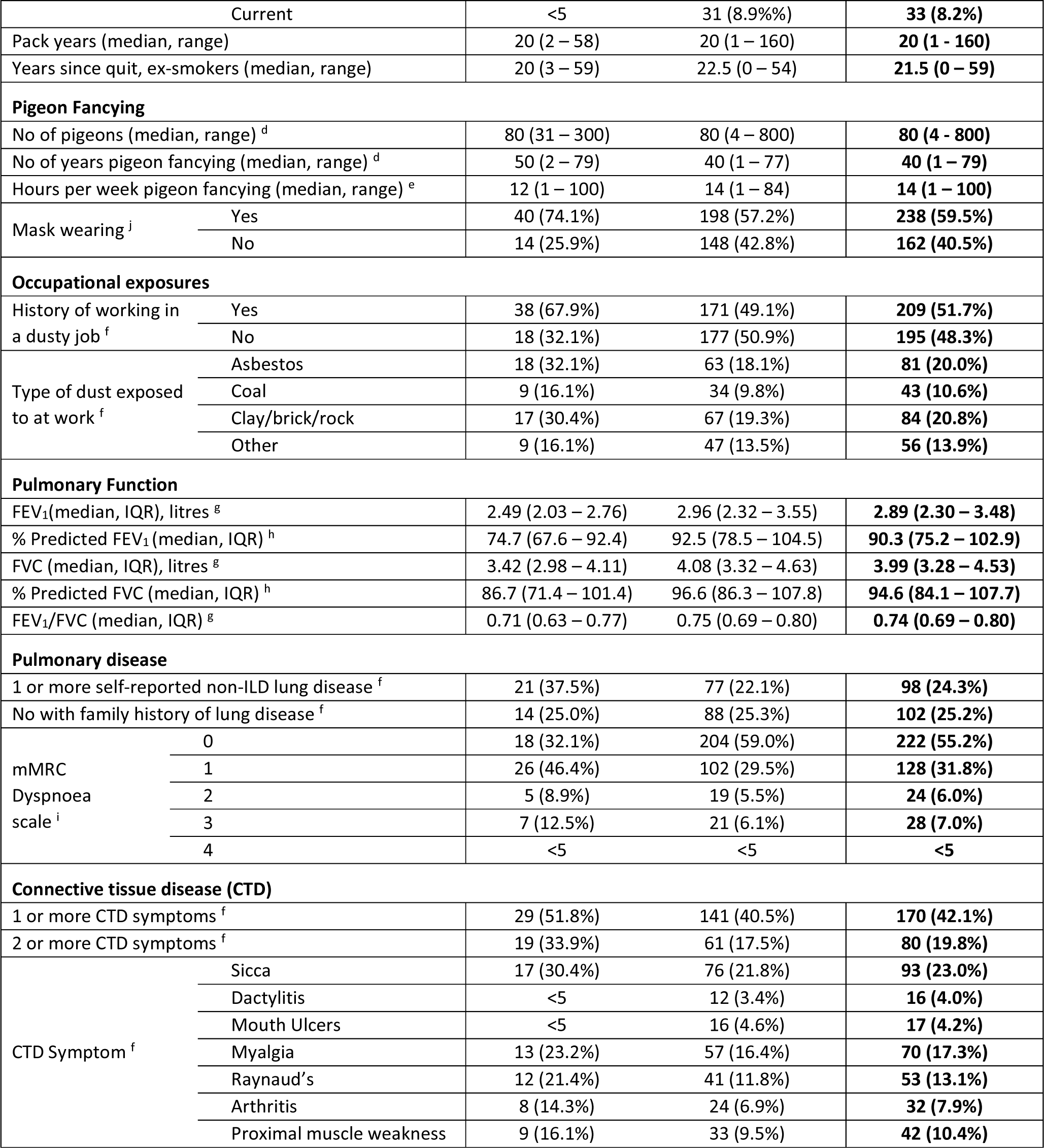
Summary of baseline study population. Baseline demographics for individuals completing a questionnaire or spirometry. Suspected interstitial lung disease (ILD) participants are defined as individuals who self-reported a diagnosis of pulmonary fibrosis or having pigeon lung and had hospital clinician review and CT scan. Percentages are presented in terms of those without missing data. n=sample size, m=metres, kg=kilograms, sd=standard deviation, FEV1=forced expiratory volume in 1 second, FVC=forced vital capacity, IQR=interquartile range. ^a^ Age and sex missing for 3 individuals. ^b^ Height and BMI missing for 33 individuals, ^c^ weight missing for 32 individuals, ^d^ 16 missing, ^e^ 15 missing, ^f^ 9 missing, ^g^ 92 missing or failed QC, ^h^ 95 missing, ^i^ 11 missing, ^j^ 13 missing, ^k^ 91 missing or failed QC.

Forty two participants volunteered a diagnosis of pulmonary fibrosis. Fifty-two participants provided a positive answer to having ‘pigeon lung’. Of these thirty-eight reported having attended a hospital clinic for assessment and having had a CT, with seven of these having had a bronchoscopy as part of their work up. Fifty-six individuals (14% of total) who reported pulmonary fibrosis or pigeon lung confirmed by CT scan were therefore defined as having a “suspected ILD” (Supplementary Figure 3).

Participants kept a median of 80 pigeons, had been involved in the sport for 40 years and spent 14 hours a week with their pigeons (Table 1, Supplementary Figure 4). Over half (60%) wore a mask when in pigeon lofts or with pigeons. A higher percentage of individuals reported wearing a mask if they had “suspected ILD” (74% in those with suspected ILD compared to 57% without).

The majority of participants had never smoked (57%), with 8% being current smokers. Around half of the study participants (52%) reported that they either currently or previously worked in an environment associated with significant levels of work-related dust. One-fifth of participants reported previous asbestos exposure via their employment. A higher percentage of individuals with “suspected ILD” reported working in a dusty environment compared to those without (68% vs 49%).

### Baseline lung health and diseases

Three hundred and twenty-one participants performed spirometry of sufficient quality (Table 1). The median percent predicted forced expiratory volume in 1 second (FEV_1_) was 90.3% (IQR 75.2-102.9) and the median percent predicted forced vital capacity (FVC) was 94.6% (IQR 84.1-107.7). Eighty-eight individuals (27% of those who performed spirometry) had obstructive spirometry (FEV_1_/FVC <0.7). Of this group twenty-nine had a pre-existing obstructive airways disease diagnosis. Individuals with “suspected ILD” had lower median measures of lung function (Supplementary Figure 5). Twenty-three individuals had measures of DLCO, and forced oscillometry measurements were performed in two hundred and twelve participants (data not presented here).

Of all individuals, 24% reported having at least one non-ILD lung disease, with the most common being asthma (n=50) and chronic obstructive pulmonary disease (COPD) (n=32). A family history of lung disease was reported by 25% of individuals, with commonest being asthma (n=30), COPD (n=21), lung cancer (n=17) and “pigeon lung” (presumed to be HP) (n=15). Individuals with “suspected ILD” had higher mMRC dyspnoea scores compared to the rest of the cohort.

Participants were asked whether they had one (or more) of ten symptoms after exposure to their pigeons (Table 2). Two hundred and one (49%) participants reported experiencing at least one symptom, and 134 (32%) experienced more than one loft-related symptom (Figure 2). The most common were wheezing (20%), sneezing (22%) and flu-like symptoms (19%). The most common symptom when in the loft was sneezing (11.6%) and the most common post-loft symptom (4 to 10 hours after being in the loft) was wheezing, reported by 12% of all participants. Around half of participants (51%) reported having at least one symptom on a persistent basis, with 20% of the participants reporting a feeling of persistent shortness of breath. Individuals with “suspected ILD” had more loft-related symptoms than those without (median of two symptoms for those with an ILD label compared to a median of zero symptoms for those without, Supplementary Figure 6).

**Figure 1:**
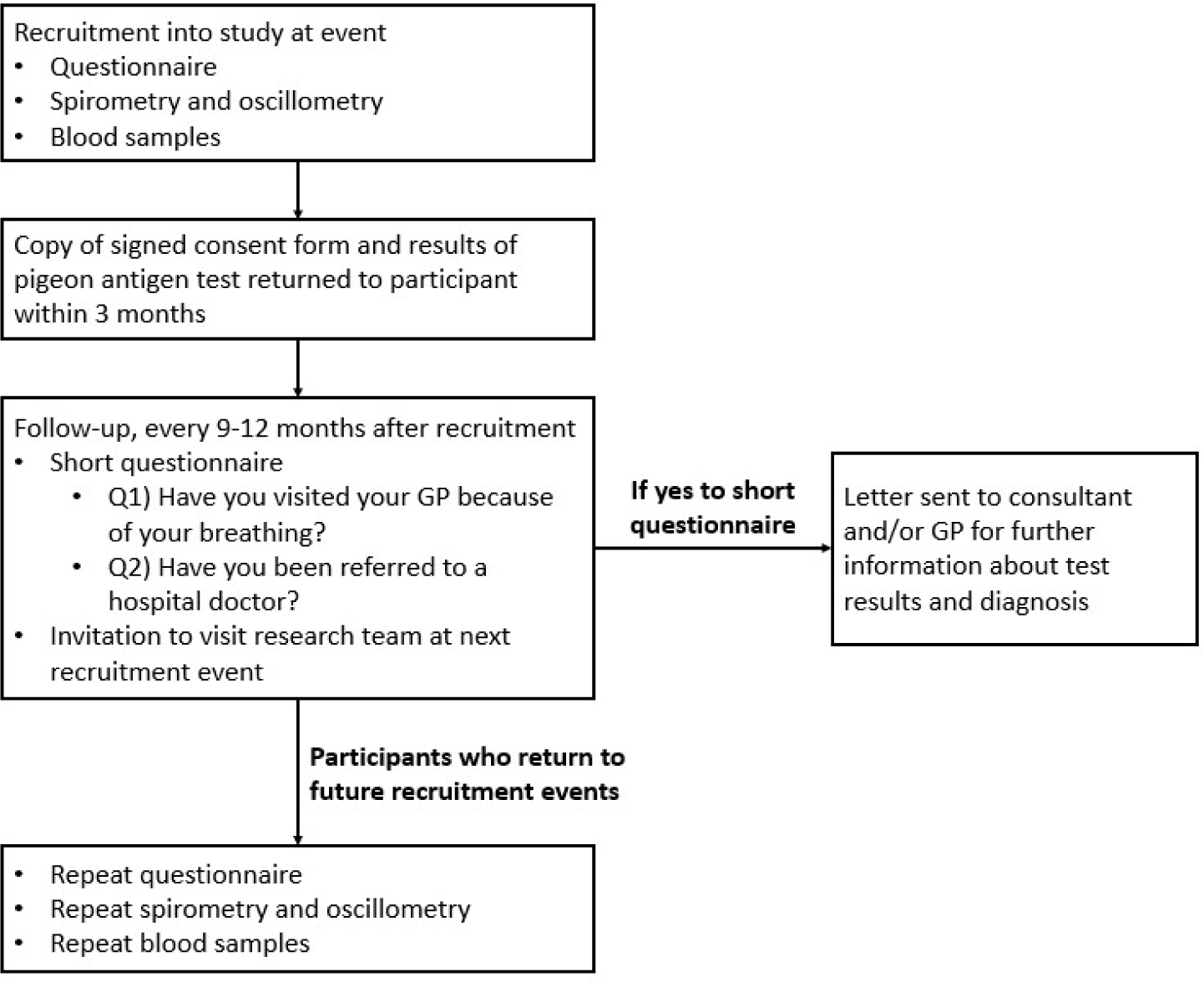
Study overview.

**Figure 2:**
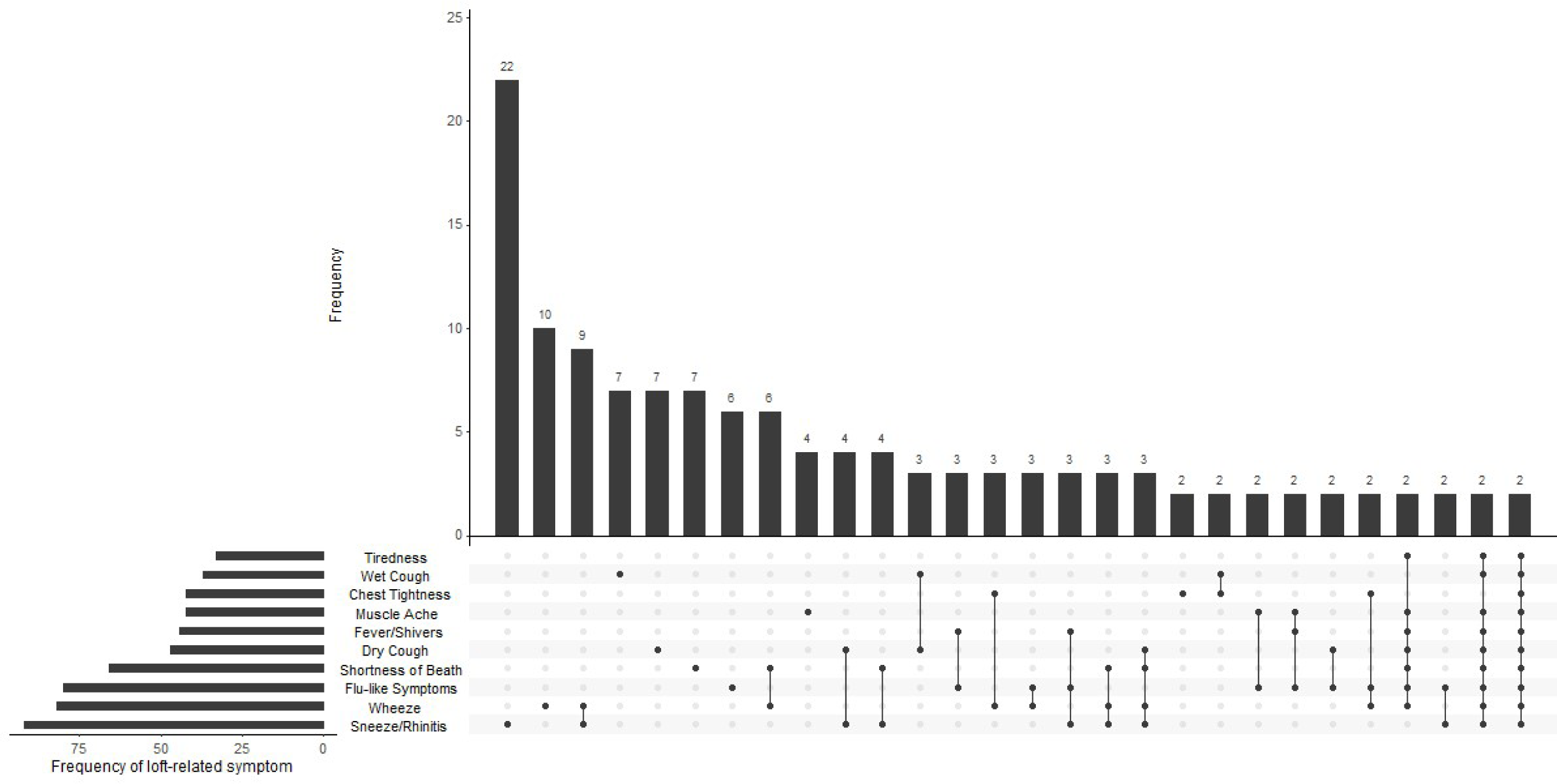
Number of loft-related symptoms. UpSet plot (9) showing frequency of symptoms associated with pigeon loft visits and which symptoms co-occur. The number of individuals with each combination of symptoms is show as vertical bars. The frequency of each symptom individually is shown by horizontal bars in the bottom left. For ease of visualisation, only symptom combinations with at least two symptoms are shown (full version of the plot can be found in Supplementary Figure 7).

**Table 2:**
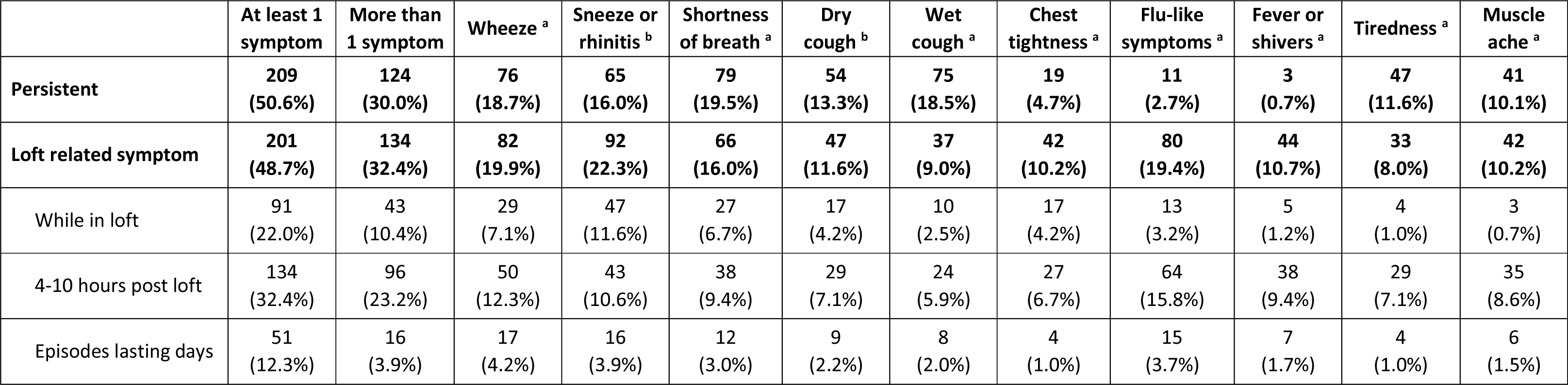
Symptoms. ^a^ 10 missing. ^b^ 11 missing. ^c^ 12 missing

|  | **At least 1 symptom** | **More than 1 symptom** | **Wheeze <sup>a</sup>** | **Sneeze or rhinitis <sup>b</sup>** | **Shortness of breath <sup>a</sup>** | **Dry cough <sup>b</sup>** | **Wet cough <sup>a</sup>** | **Chest tightness <sup>a</sup>** | **Flu-like symptoms <sup>a</sup>** | **Fever or shivers <sup>a</sup>** | **Tiredness <sup>a</sup>** | **Muscle ache <sup>a</sup>** |
| --- | --- | --- | --- | --- | --- | --- | --- | --- | --- | --- | --- | --- |
| **Persistent** | **209**<br>**(50.6%)** | **124**<br>**(30.0%)** | **76**<br>**(18.7%)** | **65**<br>**(16.0%)** | **79**<br>**(19.5%)** | **54**<br>**(13.3%)** | **75**<br>**(18.5%)** | **19**<br>**(4.7%)** | **11**<br>**(2.7%)** | **3**<br>**(0.7%)** | **47**<br>**(11.6%)** | **41**<br>**(10.1%)** |
| **Loft related symptom** | **201**<br>**(48.7%)** | **134**<br>**(32.4%)** | **82**<br>**(19.9%)** | **92**<br>**(22.3%)** | **66**<br>**(16.0%)** | **47**<br>**(11.6%)** | **37**<br>**(9.0%)** | **42**<br>**(10.2%)** | **80**<br>**(19.4%)** | **44**<br>**(10.7%)** | **33**<br>**(8.0%)** | **42**<br>**(10.2%)** |
| While in loft | 91<br>(22.0%) | 43<br>(10.4%) | 29<br>(7.1%) | 47<br>(11.6%) | 27<br>(6.7%) | 17<br>(4.2%) | 10<br>(2.5%) | 17<br>(4.2%) | 13<br>(3.2%) | 5<br>(1.2%) | 4<br>(1.0%) | 3<br>(0.7%) |
| 4-10 hours post loft | 134<br>(32.4%) | 96<br>(23.2%) | 50<br>(12.3%) | 43<br>(10.6%) | 38<br>(9.4%) | 29<br>(7.1%) | 24<br>(5.9%) | 27<br>(6.7%) | 64<br>(15.8%) | 38<br>(9.4%) | 29<br>(7.1%) | 35<br>(8.6%) |
| Episodes lasting days | 51<br>(12.3%) | 16<br>(3.9%) | 17<br>(4.2%) | 16<br>(3.9%) | 12<br>(3.0%) | 9<br>(2.2%) | 8<br>(2.0%) | 4<br>(1.0%) | 15<br>(3.7%) | 7<br>(1.7%) | 4<br>(1.0%) | 6<br>(1.5%) |

## Discussion

In this report we present the baseline characteristics of 417 individuals recruited to the BPF-GILD study. This ongoing study, which includes collection of a clinical history, breathing measurements and blood samples, along with subsequent contact to establish new relevant diagnoses in active pigeon fanciers, is ideally positioned to study the genetic and immunological determinants of acute and fibrotic hypersensitivity pneumonitis (fHP) that arise as a consequence of exposure to pigeons, and should inform subsequent work in fHP, including those cases with no identifiable cause.

A better understanding of the genetic risk factors and molecular pathways that promote fHP is required to allow the development of highly specific and unbiased tests to aid in the diagnostic process, as well as providing mechanistic insights that could lead to new treatments. Early detection and differentiation of fHP from other fibrotic lung diseases will ensure appropriate management and patient counselling, and underpin further research into the treatment of this condition. Our study, with stored blood and serum samples from a large group of well characterised subjects with regular exposure to a common driving agent and ongoing follow up, has the potential to allow detection of the relevant underlying genetic and associated pathways of relevance to the development of fHP. The spectrum of symptoms during and after exposure to pigeons described by our cohort suggest that a significant proportion are sensitised to their birds, giving us confidence we will be able to gain relevant insights.

There are some limitations to this study. Firstly, our cohort may not be fully representative of all pigeon fanciers. We have only recruited those attending either the RPRA show of the year in Blackpool, or the SHU show in Lanark, and only from those interested and willing to give up time at the meeting to complete the required assessments. In addition, at recruitment individuals were given information about measures that could be taken to reduce the risk of acute HP and related symptoms from exposure to pigeons (for example, wearing masks, improving loft ventilation and taking regular breaks from the loft). This may affect their risk of developing the condition of interest. We also recognise that individuals with higher risk of the condition could be over-represented, reflecting their being more interested in the area of research due to their symptoms or established diagnosis. Conversely under-representation may also occur, reflecting fanciers being less likely to take part as they may be concerned that they would be told to stop keeping pigeons after volunteering HP symptoms. We also only included individuals who were actively pigeon fancying, meaning individuals who had ceased fancying due to severe symptoms post exposure are not likely to be included in this study. We should also acknowledge that by using questionnaire data our results will be susceptible to recall bias and there could be an under-reporting of some exposures (such as smoking history) or symptoms. A large proportion of our cohort also report occupational exposures known to cause other forms of ILD. Finally, although the major benefit of our study is the recruitment of individuals with exposure to the same environmental antigen, results found in this study cannot be guaranteed to be generalisable to cohorts of individuals who have experienced HP associated with other exposures or undetectable causes, and any results we generate will need confirmation in these groups.

In conclusion, the BPF GILD study has recruited a large cohort of active pigeon fanciers with a spread of acute exposure symptoms. Our planned and subsequent research using data and samples obtained from this cohort is expected to result in novel findings potentially relevant to the development of fHP, which could lead to improved diagnostic accuracy and treatments for this common ILD.

## Funding and acknowledgements

The BPF GILD project was designed in collaboration with the British Pigeon Fanciers Medical Research Trust (BPFMRT) (charity SC013181), the Scottish Homing Union (SHU), and the RPRA. Funding to support the project was provided by Asthma and Lung UK, the BPFMRT, NHS Forth Valley and NHS Tayside and Wellcome Trust grant 221680/Z/20/Z. The project was sponsored NHS Forth Valley and subsequently NHS Tayside (ongoing).

## Data Availability

All data produced in the present work are contained in the manuscript

## Supplementary Methods

### Consent

Individuals were given participant information sheets about the study, with consent obtained after being allowed time to ask clarifying questions and to consider the requirements of participating in the study.

Participants were asked to provide consent for re-contact every year to allow collection of information on new diagnoses, interactions with healthcare providers, and changes to pigeon keeping activity. Participants could optionally consent to allow researchers in the study to contact their GP or respiratory consultant to obtain further information around their lung health.

### Questionnaire

Below is the questionnaire used at the Blackpool 2022 event. This questionnaire was used at all events, save the addition of a question about COVID19 and related hospitalisations from 2021 onwards.

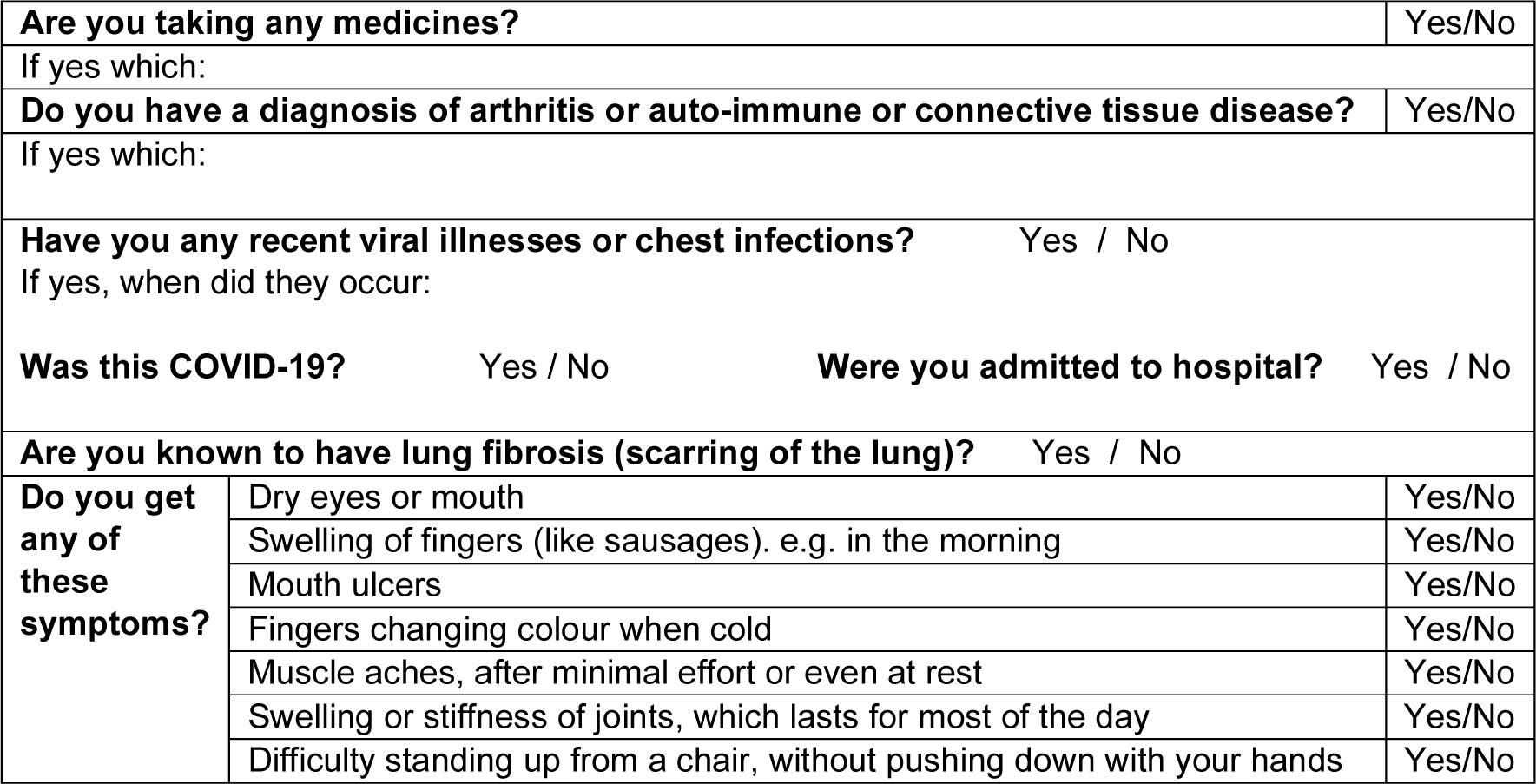

**Have you ever smoked?** Yes/No

If yes:

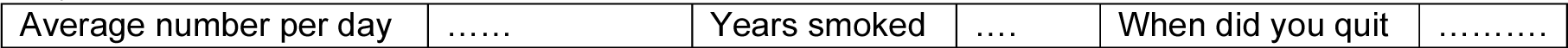

**Do/did you have a dusty occupation?** Yes/No

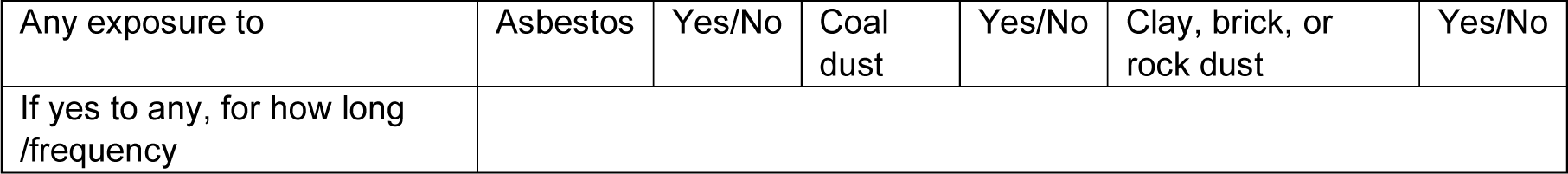

**We need to ask you how breathless you are when you are well, using the following scale (tick one):**

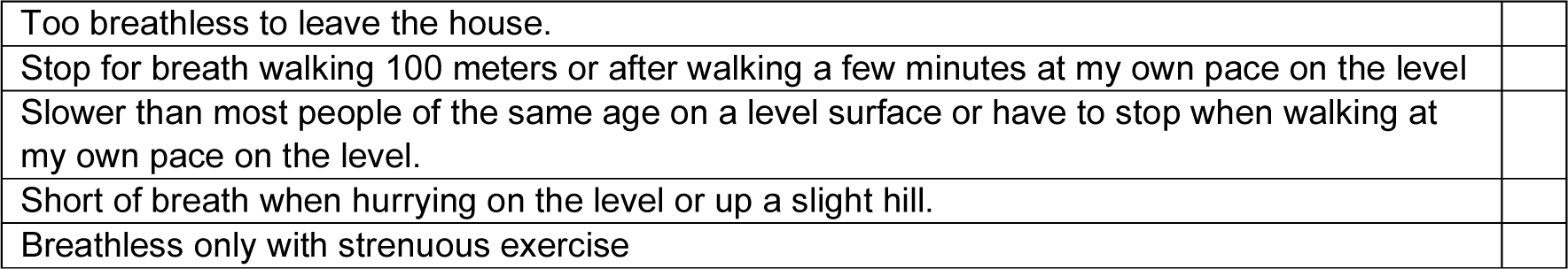

**Have you been diagnosed with ‘Pigeon Lung’ (hypersensitivity pneumonitis?)** Yes/No

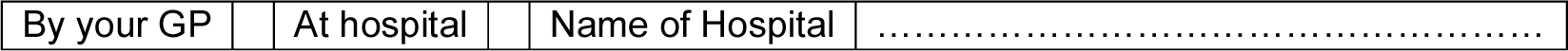

**Did you have:**

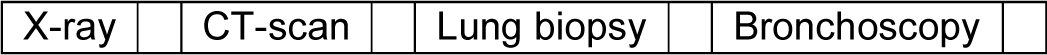

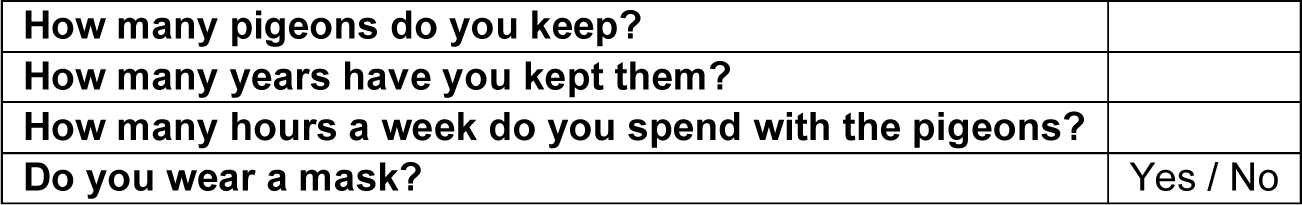

**Do you currently get any of the following symptoms (Tick any/all that apply):**

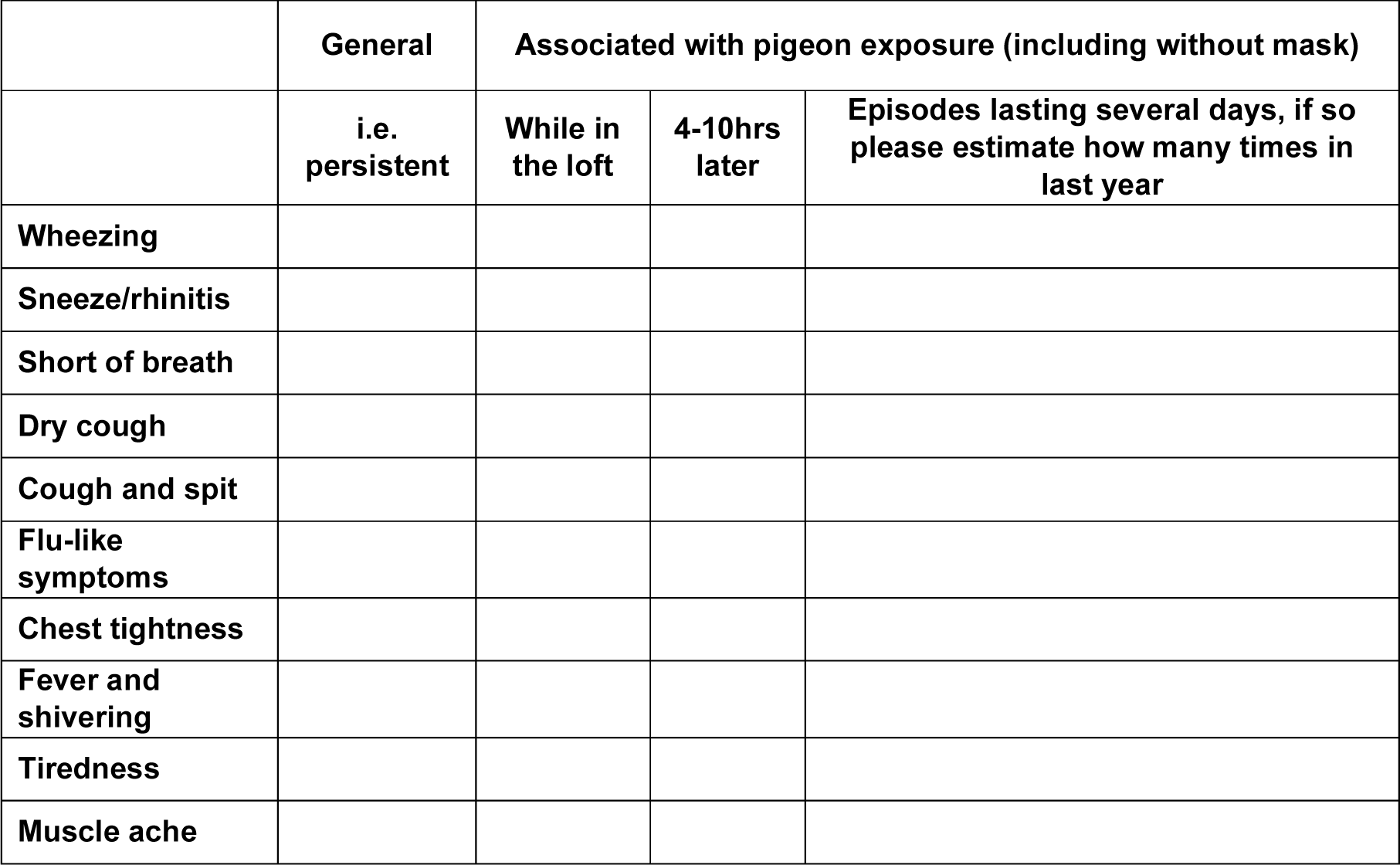

**As far as you know, has anyone in your family had a diagnosis of a lung condition?** Yes/No

**If yes, what do you remember it being called?**

**Have you ever had a diagnosis of any other lung disease?**Yes/No

**If so, what name has it been given?**

---

**Supplementary Figure 1:**
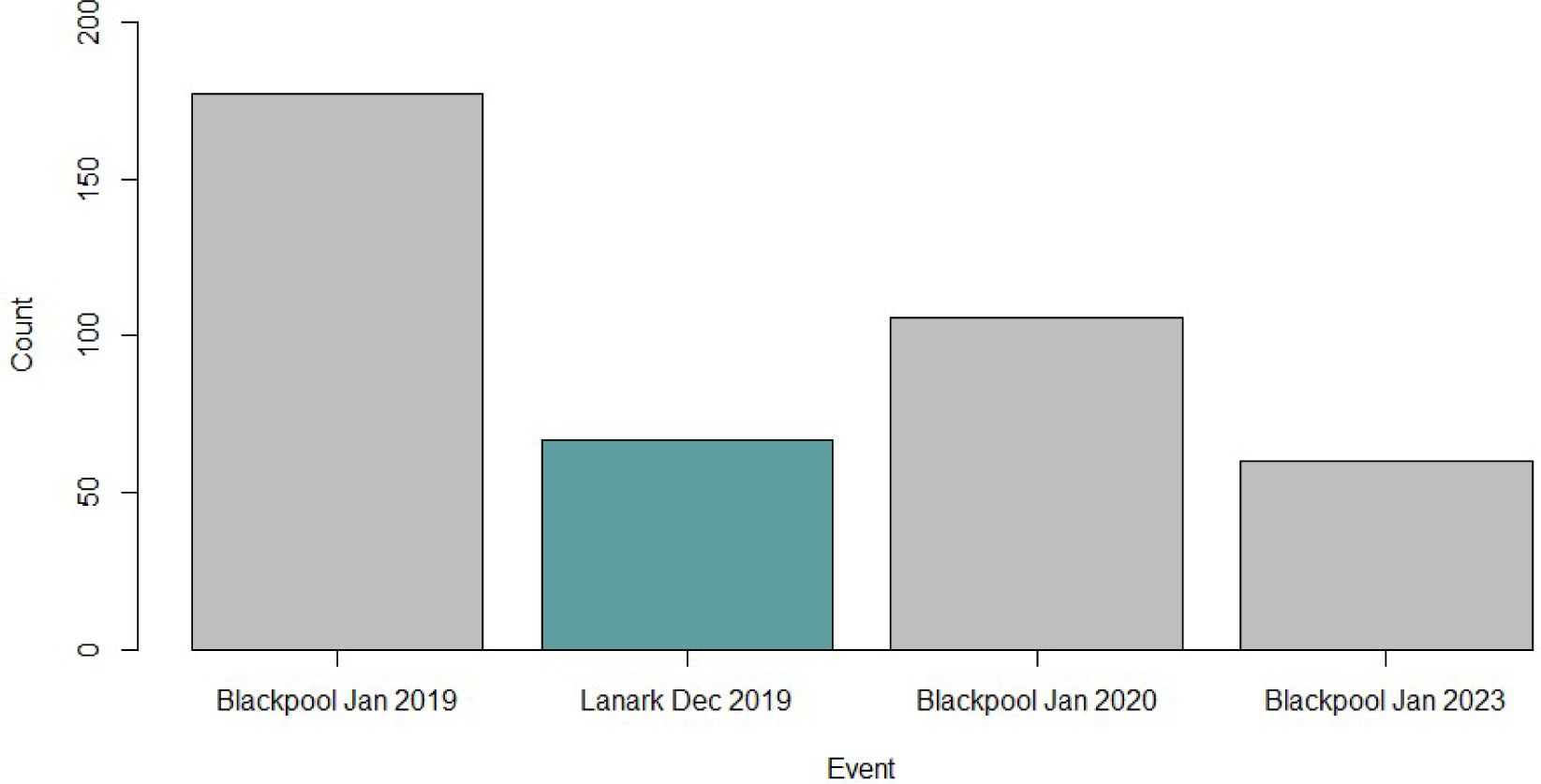
Recruitment event. The bar plot shows the event that individuals were first recruited to the study.

**Supplementary Figure 2:**
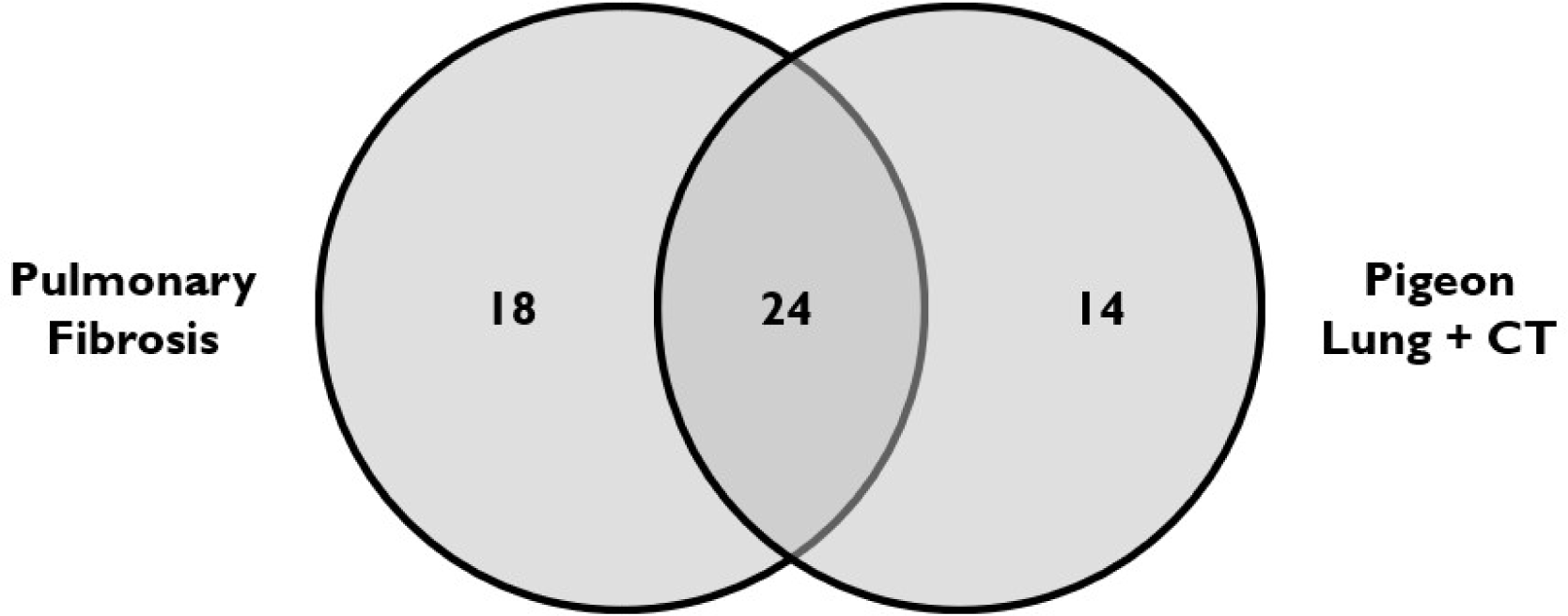
Overlap between participants reporting pulmonary fibrosis and pigeon lung (with hospital clinic review and CT scan). The 56 individuals here who either self-reported pulmonary fibrosis or “pigeon lung” (with hospital review including CT) were defined as having “suspected ILD”. There were 24 individuals who self-reported having both pulmonary fibrosis and pigeon lung.

**Supplementary Figure 3:**
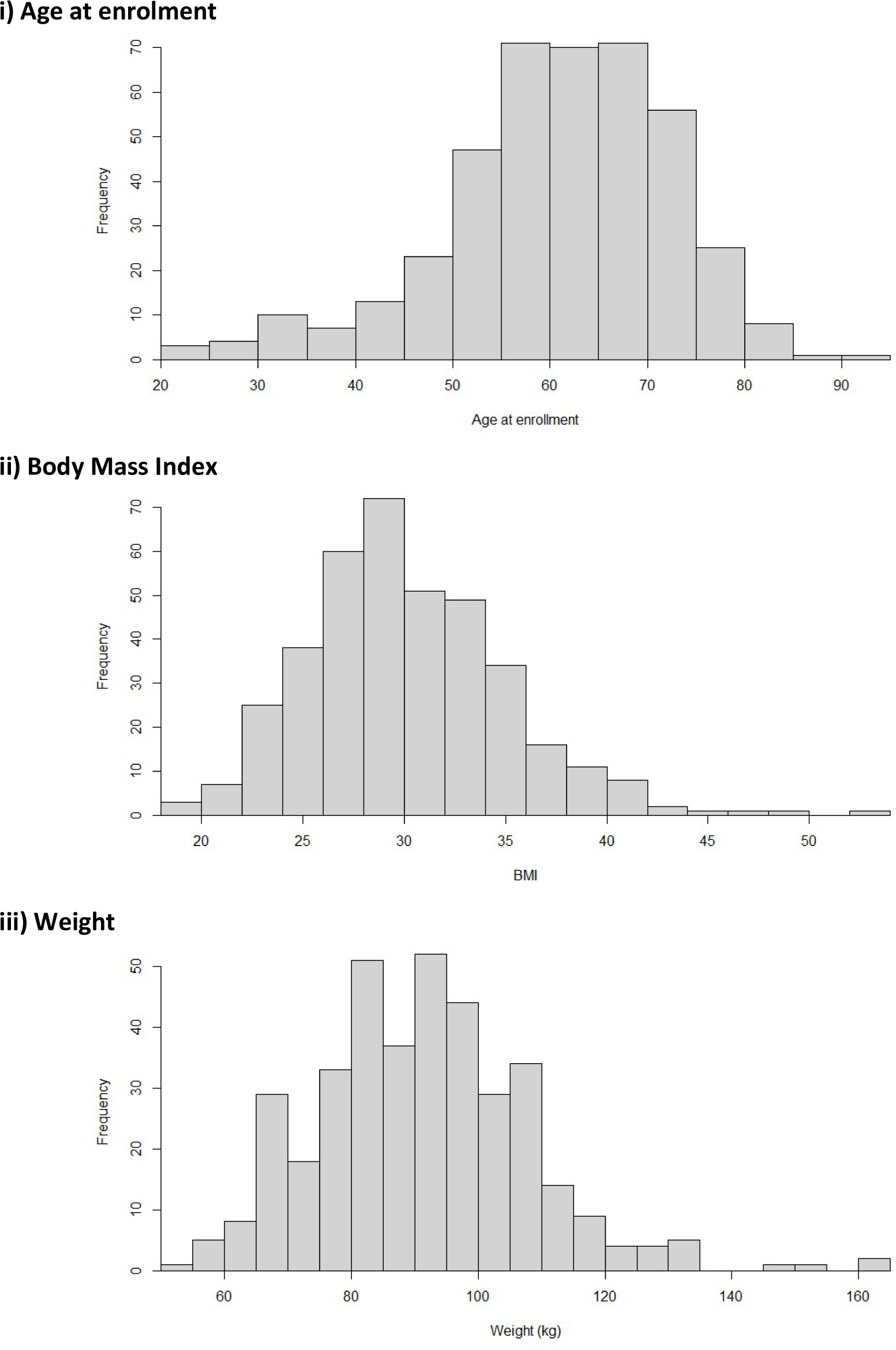
Histograms of baseline demographics.

**Supplementary Figure 4:**
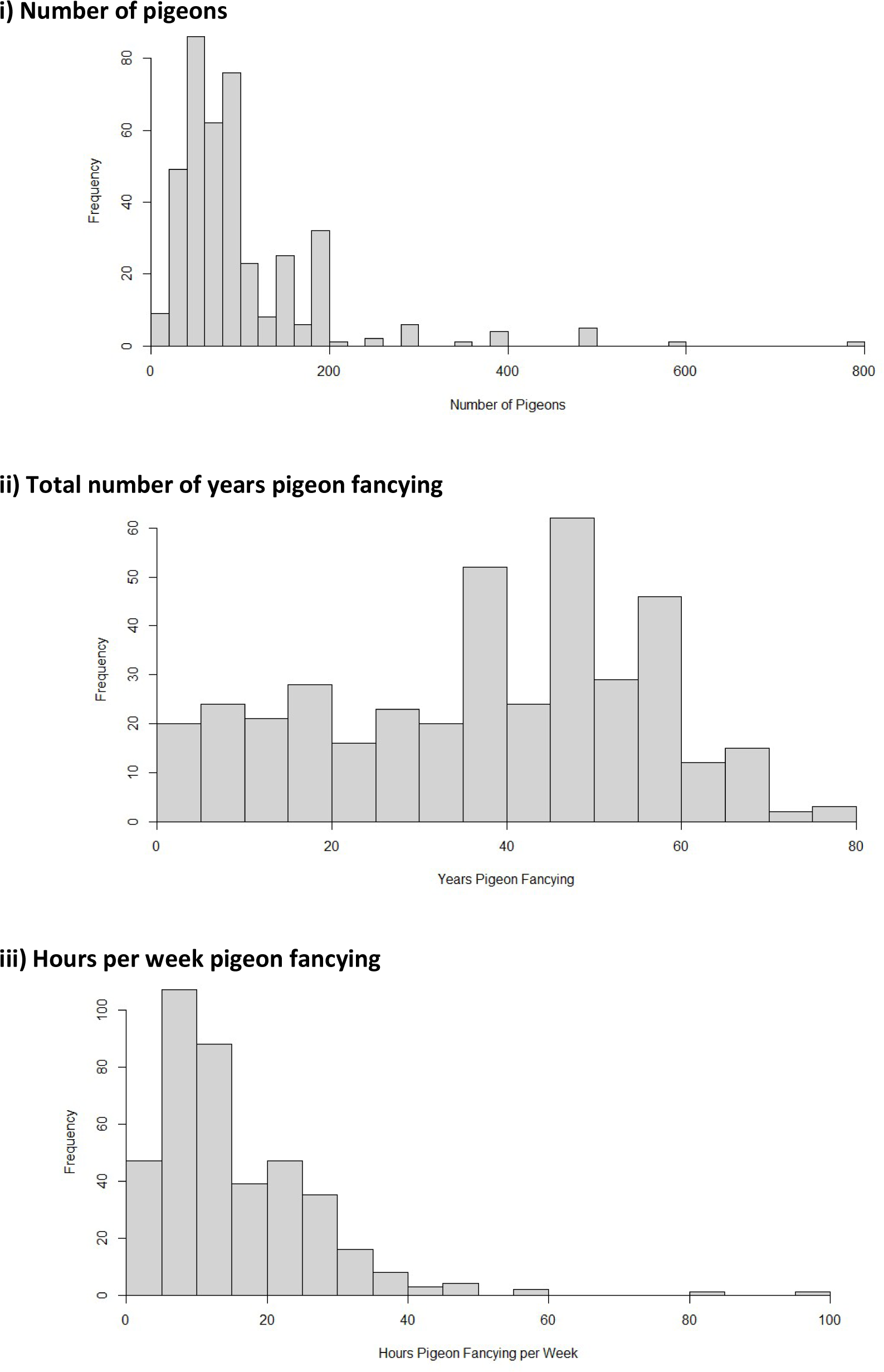
Histograms of pigeon keeping activity.

**Supplementary Figure 5:**
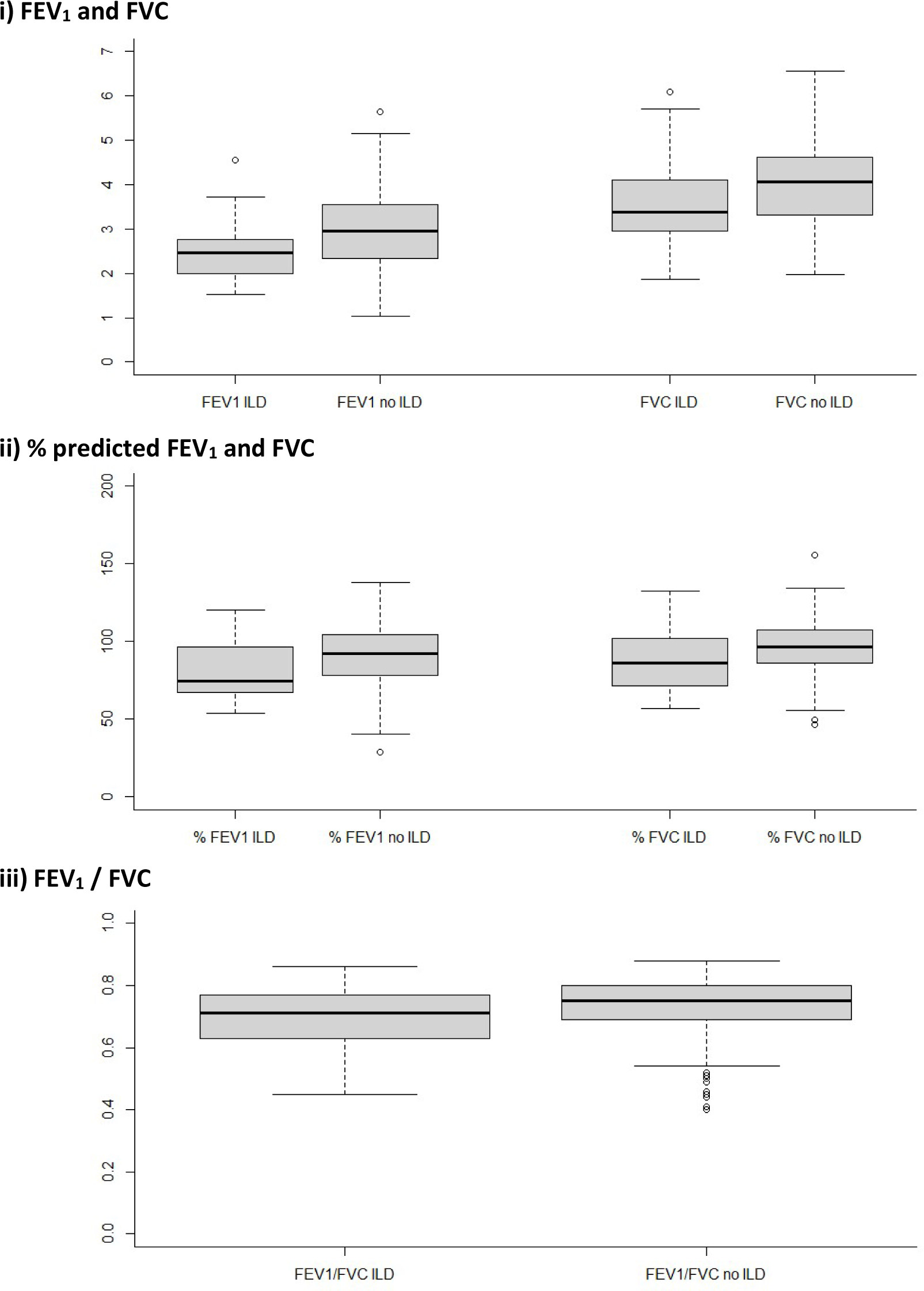
Box and whisker plots of spirometry. Results are split by whether individuals had “suspected ILD”. FEV_1_: Forced Expiratory Volume in 1 second, FVC: Forced Vital Capacity.

**Supplementary Figure 6:**
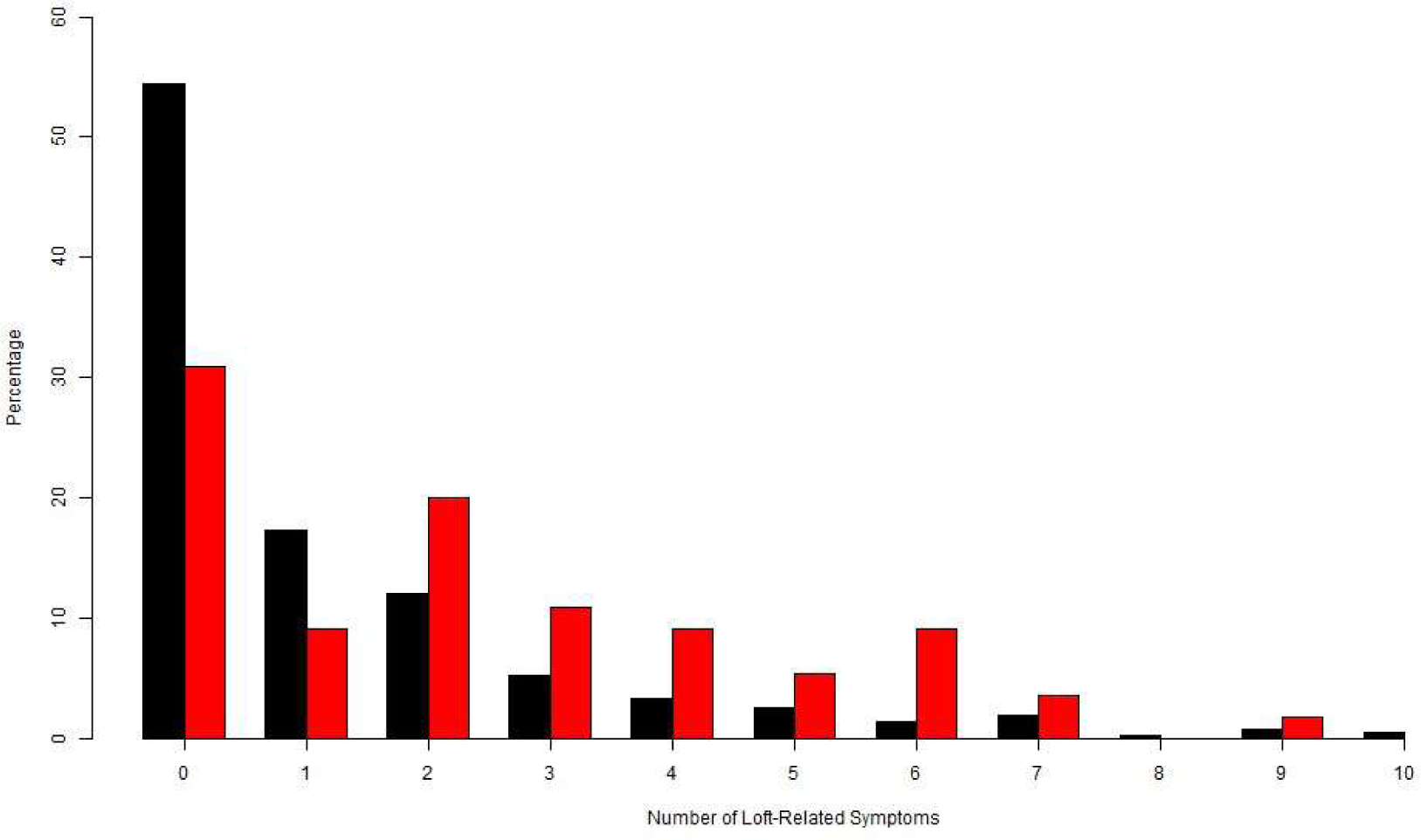
Number of loft-related symptoms. Bar plot showing the number of symptoms reported by participants when in loft or following visit. Individuals with a suspected ILD (i.e. defined as self-reporting diagnosis of pulmonary fibrosis or a diagnosis of ‘pigeon lung’ with associated with hospital clinician review and CT scan) are shown in red, individuals without a suspected ILD are shown in black.

**Supplementary Figure 7:**
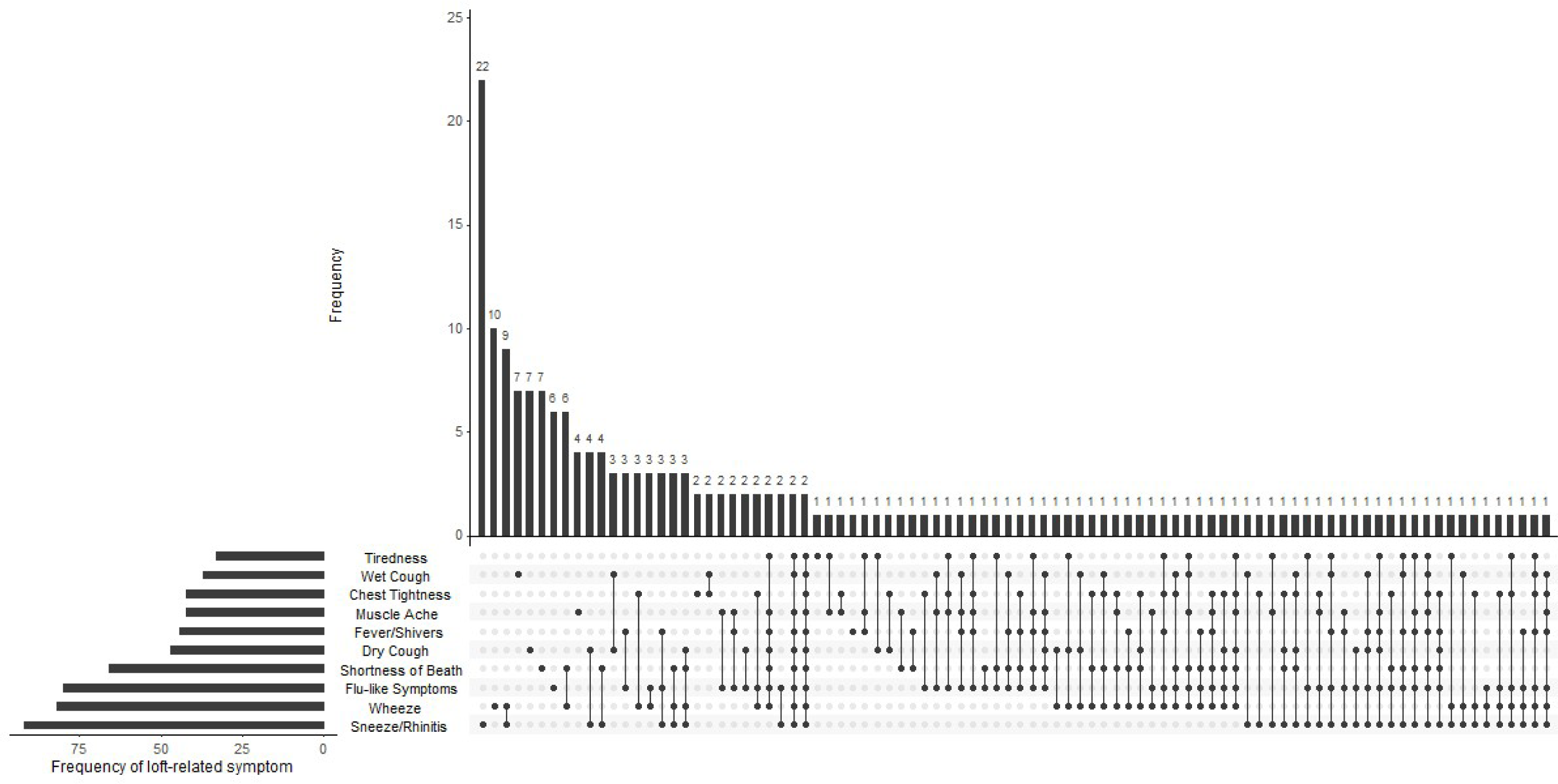
‘UpSet’ plot showing combinations of loft related symptoms.

